# Social, behavioural, and cultural factors of HIV in Malawi: a semi-automated systematic review

**DOI:** 10.1101/2020.03.12.20034702

**Authors:** Amaury Thiabaud, Isotta Triulzi, Erol Orel, Kali Tal, Olivia Keiser

## Abstract

**Background:** Demographic and socio-behavioural factors are strong drivers of HIV infection rates in sub-Saharan Africa. These factors are often studied in qualitative research but ignored in quantitative analyses. However, they provide an in-depth insight into the local behaviour, and may help to improve HIV prevention.

**Methods:** To obtain a comprehensive overview of the socio-behavioural factors influencing HIV prevalence and incidence in Malawi, we systematically reviewed the literature. Due to the choice of broad search terms (“HIV AND Malawi”), our preliminary search revealed many thousands of articles. We, therefore, developed a Python tool to automatically extract, process, and categorise open-access articles published from January 1st, 1987 until October 1st, 2019 in Pubmed, Pubmed Central, JSTOR, Paperity, and arXiV databases. We then used a topic modelling algorithm to classify and identify publications of interest.

**Results:** Our tool extracted 22’709 unique articles; 16’942 could be further processed. After topic modelling, 519 of these were clustered into relevant topics; 20 of which were kept after hand-screening. We retrieved 7 more publications after examining references so that 27 publications were finally included in the review. Reducing the 16’942 articles to 519 potentially relevant ones by using the software took 5 days. Several factors were identified to contribute to the risk of HIV infection, including religion, gender and relationship dynamics, beliefs, and socio-behavioural attitudes.

**Conclusions:** Our software does not replace traditional systematic reviews, but it returns useful results to broad queries of open-access literature in under a week, without a priori knowledge. This produces a “seed data-set” of relevance which could be further developed. It identified known factors and rare factors that may be specific to Malawi. In the future, we aim to expand the tool by adding more social science databases and applying it to other sub-Saharan African countries.

## Introduction

Demographic and socio-behavioural factors are strong drivers of HIV in sub-Saharan Africa, but the interactions between these factors—the way their influence shifts over time and space and influences HIV prevalence and incidence—are poorly understood. Some epidemiological studies reported on spatial variability of the HIV epidemic, using statistical analyses to assess the association between the spatial distribution of HIV prevalence and potential risk factors [1-7]. They found, for example, that high population density [7] or a short distance to a road or a clinic [6] were associated with a high HIV prevalence. Tomita et al. showed that behaviour (sexual debut, uptake of contraception and circumcision) and social determinants strongly influenced the risk of HIV acquisition [8]. An analysis of 29 sub-Saharan African countries found associations between 12 demographic and socio-behavioural factors, including variables related to age, literacy, HIV knowledge, domestic violence, women’s empowerment, and sexual activity [9]. The patterns of associations were complex and varied by sex and country, but the study did not include many potentially significant factors because they were absent from the data or were only available for some countries and it did not consider subnational variation. These epidemiological studies did not draw on qualitative research and they rarely contextualized the associations they identified.

Social scientists of various disciplines have performed qualitative studies of social and cultural factors related to HIV, providing rich detail on the perceptions and behaviours of people in specific localities. For example, medical anthropologists have examined maternal care-seeking behaviour in different geographic regions and groups [10]. Cultural studies have analysed connections between belief in witchcraft and folk epidemiological wisdom about HIV [11], and the connection between women’s educational level and attendance at antenatal care [12]. Sociologists studied the perceptions of HIV testing in rural Malawi [13], or have analysed power structures [14] and the vulnerability of adolescent girls [15].

Qualitative research gives us an in-depth understanding of local situations and may help identify factors that quantitative analyses have not considered. Qualitative analyses often focus on individual knowledge, opinions, attitudes and challenges, while quantitative analyses quantify relationships between various factors and between factors and outcome variables. Combining qualitative and quantitative studies may reveal how and why various factors interact across time and space in a complex and widespread epidemic.

We used the topic of HIV in Malawi as a case study for an in-depth literature review of quantitative and qualitative literature on social and behavioural factors that may influence the HIV epidemic. We designed the review to be broad and inclusive to capture all possibly relevant factors and expected it to identify known factors, neglected factors, and some factors that may never have been identified or analyzed in quantitative studies. We chose Malawi because 1) Malawi has relatively high HIV prevalence that varies substantially between regions [16]; 2) the country is socio-culturally diverse because it is home to many ethnic groups (e.g., Chewa, Nyanja, Tumbuka, Yao, Lomwe, Sena, Tonga, Ngoni, Ngonde, Asians, and Europeans); 3) a preliminary search revealed that Malawi was the focus of many scientific studies on HIV, including some by our group [17-19].

Because we knew that the breadth of our topic would return too many studies to read, we developed a semi-automated literature search engine and software that automatically downloads and analyzes open access full-text articles; this software can be used for searches on any broad topic across any regions.

## Methods

### Search strategy

We searched all English language articles published from the inception of the databases from January 1st, 1987 until October 1st, 2019 using the query “HIV AND Malawi”. We performed an automated search of Pubmed (PM), Pubmed Central (PMC), Paperity (PAP), and arXiV, using a custom Python script and the corresponding application programming interfaces (APIs). We also sent a request for the same type of data to JSTOR since it offered no API to directly access the database.

### Inclusion and exclusion criteria

We used a broad query designed to capture all articles about HIV and Malawi, not just those focused on health, and considered all studies that discussed social, behavioural, and cultural factors that might be associated with HIV infection in Malawi. The selection process is described in more detail below. We included original peer-reviewed articles, both quantitative and qualitative studies, and preprints. We also analysed systematic reviews but preferred to include original publications when possible. We discarded articles that investigated the effects of HIV/AIDS (e.g., [20]).

### Data collection

Figure 1 shows the pipeline along which data were collected and processed. We extracted the following information from articles: list of authors and their affiliations; MesH keywords; Digital Object Identifier (DOI); title; abstract; full-text; publication date; journal provider; and URL of the PDF version of the full text article. Mandatory fields were title and availability of the full text; other items were retrieved if possible. If full-text articles could not be directly retrieved from the database, they were extracted from either an automatically downloaded PDF when the URL of the PDF was available or retrieved through an automatic reversed search with the DOI resolver [21]. The DOI resolver identified PDFs by scraping the internet. In some cases, the information in databases was incomplete (e.g., missing DOI) or we could not access the PDF (e.g., access restrictions, PDF contained only images, unavailable PDF) so we could not obtain the full text. We then checked the data for duplicates and merged them.

**Figure 1.**
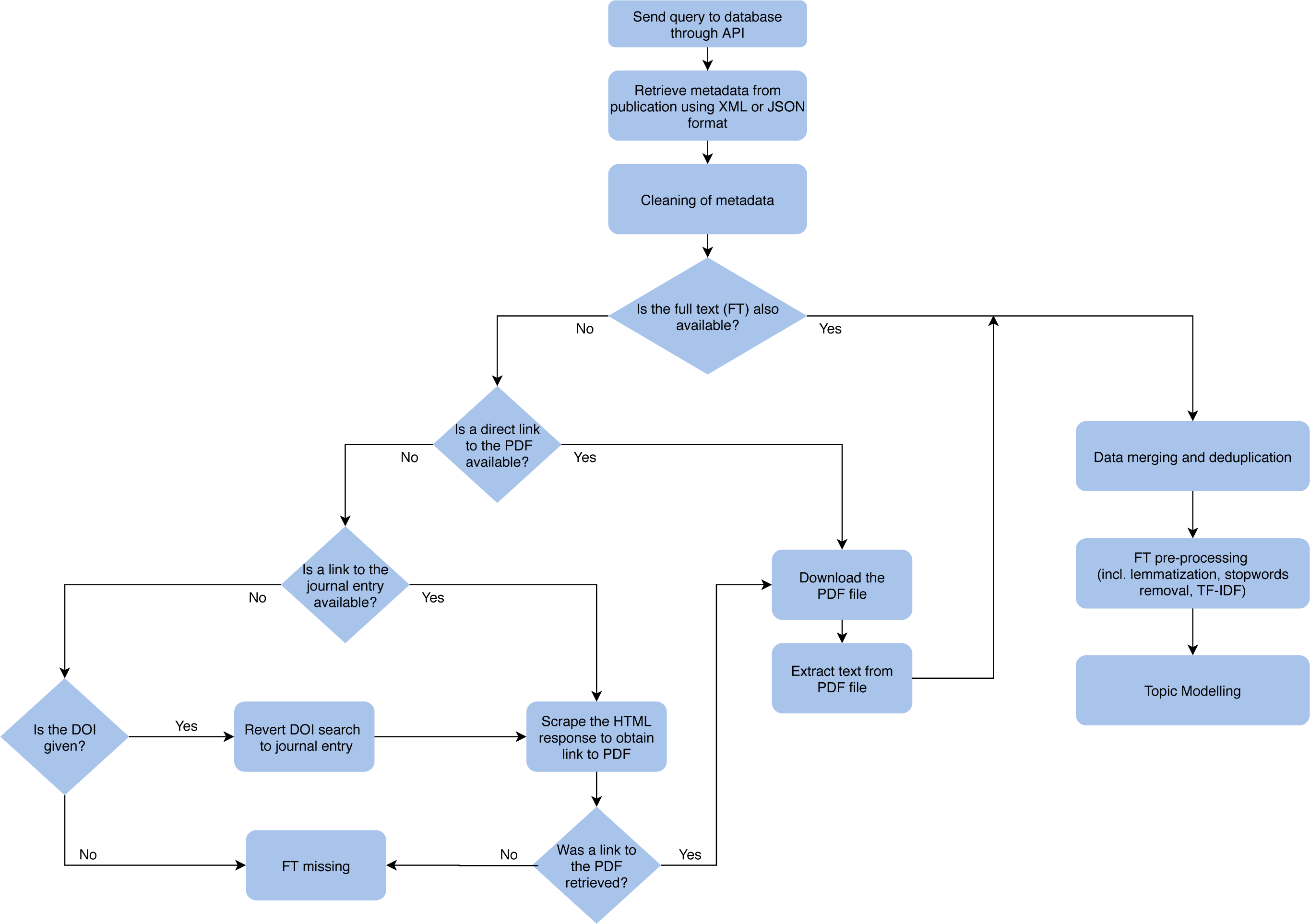
Pipeline schematics for gathering and preprocessing data.

Each extracted full-text article was tokenized and processed further. We deleted non-ASCII characters, numbers, words under 4 characters (except some acronyms of interest like “HIV” and the abbreviation for antiretroviral therapy, “ART”), and a list of stopwords from the NLTK Python toolkit [22] with relevant additions (e.g. URL, “volume”, “journal”). We then lemmatized the text to avoid duplicating words with different inflectional endings.

Once cleaned, we ran frequency-inverse document frequency (TF-IDF) from the scikit-learn Python package to extract relevant keywords [23]. Because the formatting of author affiliations was so heterogeneous, we retained only the city. We then stored original data and the generated data in a local SQLite database.

### Topic Modelling

Finally, we classified documents into topics based on their similarity with topic modelling. The process allowed us to broaden our search terms to identify relevant publications and to extract keywords relevant to a topic, providing an overview of salient and relevant terms that may best represent the data. We scored each document for a set number of topics, using the Latent Dirichlet Allocation (LDA) method [24]. Based on their highest topic score, publications were allocated to a topic.

To optimize computational efficiency and quickly identify potentially relevant articles, we initially chose five topics; for each, we used the same approach to identify five subtopics. We repeated the process four times and identified 625 topics. The resulting “tree of topics” increased its specificity at each repetition, which helped us identify our topics of interest faster than if we had started with many topics. We manually selected the number of iterations, but our selection was analysis-driven. Although more iterations would have reduced the number of potentially relevant articles that we needed to check manually, it would have raised the risk of missing relevant articles, and the performance of the algorithm would have decreased.

## Results

The PRISMA diagram (Figure 2) summarizes the selection process for relevant articles. Based on our search term, the software extracted a total of 22’709 unique articles, of which 16’942 full texts (74.6%) were retrieved directly from the database or extracted from PDFs. Topic modelling automatically screened these articles, reducing our selection to a subset of 519 relevant articles that included 14 topics related to behaviours, beliefs, culture, and religion. Of these, we manually selected 119 based on titles and abstracts and after applying our inclusion and exclusion criteria. After full-text screening, 20 articles were included in the systematic review. We identified 10 more articles from references and included 7 of them. The systematic review finally included 27 publications: 20 (74.1%) were quantitative, and 7 (25.9%) were qualitative. It took about 5 days to retrieve and preprocess the data for topic modelling. The iterative topic modelling took about a day.

**Figure 2.**
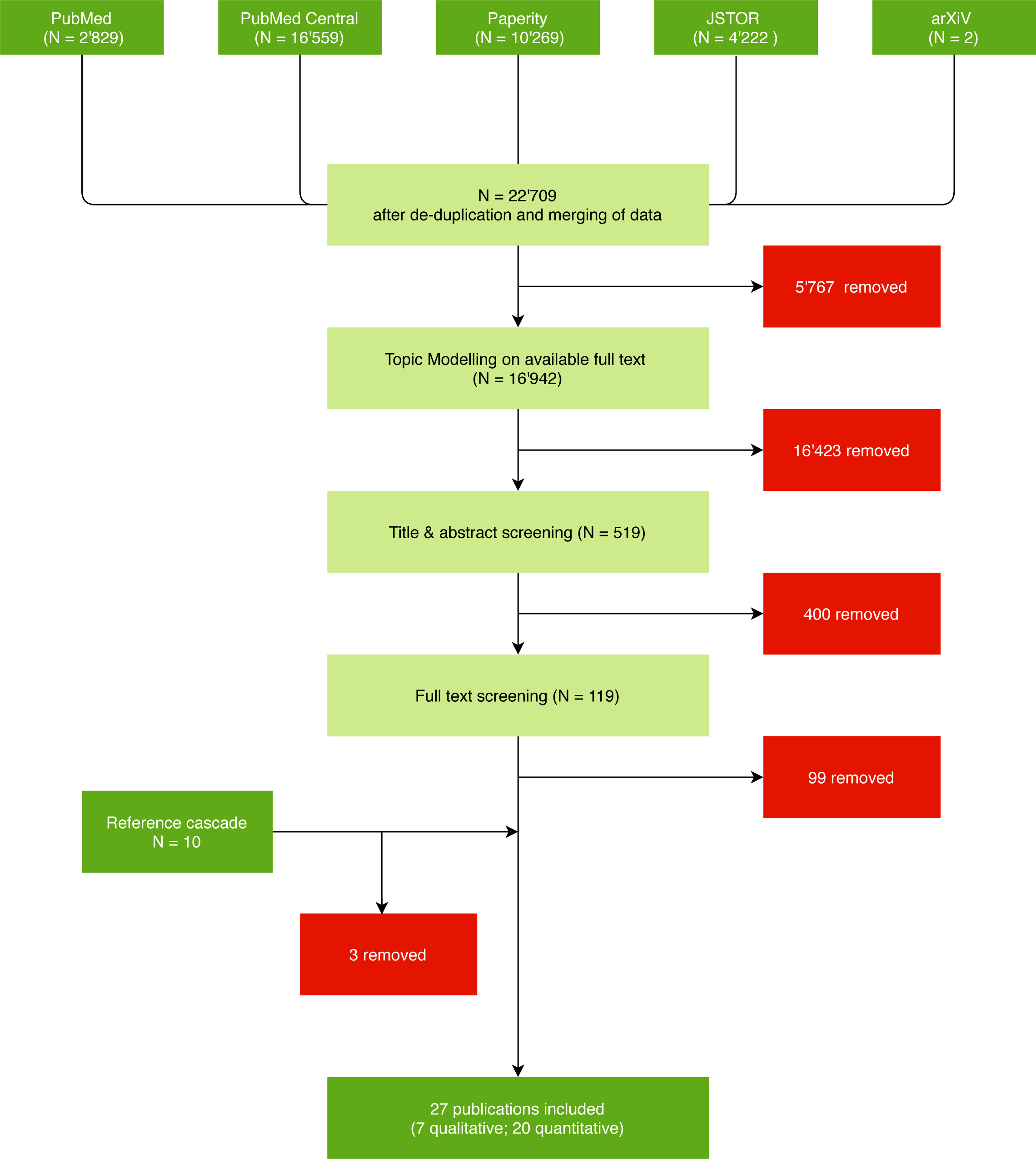
Study selection.

Below, we summarize the main findings of the included articles by topic area.

### Religion

We identified three publications that found religion was associated with the risk of HIV transmission or decreased risk behaviour. All noted the effect of the religious leader on family planning, sexual morality, positive attitude towards condom use [25], and reducing stigma [26,27]. The studies also showed that congregations in rural Malawi responded to the HIV epidemic by taking care of orphans, helping the sick, or sponsoring HIV-related knowledge programs [26,27]. In this way, religion shaped the HIV experience of congregation members. Yeatman et al. [25] noticed an association between religious socialisation and contraceptive use. The literature search also highlighted that Muslim men were more likely to be circumcised, and less likely to be HIV infected than men of other faiths [24], though they were also less likely to use condoms. Being religious did not necessarily help prevent HIV, though Lau et al. [28] noticed a positive association between weekly attendance of religious services and women’s use of modern contraceptives. Faith was often associated with condom rejection. Anglewicz et al. [29] found that Muslim men tended to find condom use less acceptable than did those of other faiths, while a change to Christianity increased condom use in men.

### Partnerships and gender

Reniers et al. [30] investigated the role of polygyny in Rumphi, Mchinji and Balaka. They reported more extramarital sex in polygynous marriages than in monogamous marriages. They further found evidence that in polygynous marriages, the latest wives were more likely HIV-positive than the first wives. Other studies also showed that having more than one sexual partner increased HIV prevalence, led to differences in adherence counselling and testing, or differences in male circumcision practices [28,31,32]. Stephenson et al. [33] found a discrepancy between different types of relationships: cohabiting men were less likely to have risky extramarital sex than married men.

Age-asymmetry in relationships plays a role in HIV transmission [34]. One article examined the age-difference between women and their partners on HIV transmission on Likoma Island. When male partners were 2 to 12 years older than their female partners, the women’s risk of being HIV-positive was higher than in women whose partners were more than 12 years older. When female partners were more than 5 years older than their male partner, they were more likely to be HIV-positive than women whose partners were 0-4 years older. Never using condoms and being married were associated with larger age differences between men and women.

Condom use helps prevent HIV infection. Anglewicz et al. [29] showed that marital status and women’s and men’s risk perceptions were associated with condom use. Getting married reduced the acceptability of using condoms during sexual intercourse. A woman’s perception of her HIV status was generally more important than her real HIV status for the acceptability of condom use within marriage. Known HIV status was a more important determinant of condom use in men than in women [29].

Power structures and closeness in partnerships may strongly influence HIV transmission. Becker et al. [31] showed that in women, prior HIV testing and emotional closeness to a partner were associated with acceptance of home-based HIV testing and counselling services. Schatz showed that married women may be able to protect themselves from infection by communicating with their husband about HIV, by confronting their husband’s sexual partners, or by refusing polygyny. Women may also seek support and advice from their friends and relatives, or even ask for a divorce. This suggests that women are not always vulnerable; some can self-advocate for their protection in marriage to reduce their risk of acquiring HIV, especially in the matrilocal southern part of Malawi where a woman can tell her husband to “take your mat and go” [35]. Even in the patrilocal northern region, Schatz [35] highlights the support of women by their kin if the husband’s risky behaviours cause women to return to their family.

### Beliefs

We found several articles that studied beliefs and found associations between misconceptions and ignorance about HIV transmission factors and risky behaviour. Authors labelled these “false beliefs” and found they stigmatized people with HIV [25,36,37]. One study mentioned beliefs about women’s cleansing rituals. Some congregations believe women will be cleansed by having unprotected sex (e.g. after the death of their husbands, after giving birth, or after miscarriage) [38]. The belief that HIV-infected men can be cured by having sex with a virgin woman also spreads the disease.

Personal beliefs play an important role in HIV prevention and risk of HIV infection. Three studies highlighted an important role of perceived HIV status between partners. Anglewicz et al. [39] examined the accuracy of perceived HIV status in 768 monogamous couples, finding that partners tended to overestimate the risk of being HIV-positive; overestimation was associated with marital infidelity. But knowing one’s or one’s spouse’s actual HIV status significantly reduced HIV risk. Fedor et al. [40] showed that once HIV-negative women and HIV-positive men learn their status they reduced risky behaviour by increasing condom use and having sex with fewer partners.

### Social and Behaviour

We identified three studies on the effect of behaviour-change interventions on HIV [41-43]. In two studies [41,43], they found that the intervention seemed to affect HIV risk behaviours and knowledge; the third found none [42]. Crittenden et al. [41] studied the spread of behavioural and psychological factors with peer group interventions in central Malawian adults living in rural areas. The behavioural changes that were promoted in the intervention group (e.g. partner communication, use of condoms, recent HIV test) spread to other persons in the same community. The second study [43] assessed the effect of a cash transfer program (lottery ranging from US$1 to US$ 5) in adolescents and women aged 13-22 years who attended school. The primary outcome was HIV prevalence 18 months after study enrollment; it was 1.2% in the intervention group and 3.0% in the control group. Women who received the cash transfer were less sexually active than women in the control group. In contrast, a study in Northern Malawi [42] showed that behaviour change interventions did not reduce risk of HIV infection in Malawian adolescents, possibly because these interventions send contradictory messages or because adolescents are more influenced by their living environment (culture, religion, peers).

Several studies investigated the association between migration and HIV. Helleringer et al. [44] reported an association between concurrent partnerships and HIV serodiscordance among couples on Likoma Island. HIV positivity was associated with migration out of the country (circular out-migration) and sexual contact with temporary in-migrants to the island. Anglewicz et al. [45] concluded that migrants were more at risk for HIV infection but migration was not the reason for the higher risk: people with HIV were more likely to migrate, thereby reversing the causality.

Low socioeconomic status often drives HIV in Malawi [38,43]. It was associated with early sexual relationships, transactional sex, and a higher probability of having sex with older men [38]. In some studies, a low socioeconomic status was also associated with coercive heterosexual relationships. Coercive sex was a strong predictor for HIV infection in male victims [46]: the likelihood of being HIV positive was 7.2 times higher among men who had been sexually coerced than among those who had not been. One publication studied the association of coercive sexual behaviour with social and economical status [47] and found that unemployment was strongly associated with coercive sex in young men in Blantyre, whereas material deprivation only was strongly associated with coercive sex in young women.

HIV acquisition was associated with intravaginal practices and products applied by women to manage their sexual relationship, menstruation, and to improve their health. Women used cloth or paper to wipe out their vagina, they inserted products to dry or tighten the vagina, and they used intravaginal cleaning soap [38,48]. The use of the injectable hormonal birth control drug medroxyprogesterone acetate was associated with HIV seroconversion in HIV-negative women during a clinical trial [32]. Among men who have sex with men (MSM), the use of water-based lubricants could lower HIV risk [49]. Being older than 25 years, not being married, and age at first sexual intercourse were associated with HIV infection [49]. Some of these variables were also identified in other studies [28,31,46].

Although studies from other sub-Saharan African countries showed an association between alcohol and drug consumption with HIV testing, HIV infection, or uptake of preventive methods, evidence was limited in our study. Lau et al. [28] found no association between tobacco use and male circumcision. Conroy et al. [64] found some association between alcohol use and being HIV positive, but the association was not statistically significant (odds ratio 1.56). They did find a significant association between sexual coercion of women and alcohol use among men.

## Discussion

### Strengths

Using a broadly inclusive search phrase and repeated topic modelling we quickly identified a small number of highly relevant articles about socio-behaviour factors and HIV in Malawi among 16’942 open access articles from five different databases. Our Python tool quickly reduced the number of potentially relevant articles to 519 in a few hours. It took us 5 days to screen titles and abstracts of these 519 articles, identify 119 potentially full-text articles, and include 20 remaining articles in the review. We then added 7 more articles from references.

Our software allowed us to omit the time-consuming step of devising and tuning a specific search query combining logic keywords (« AND », « OR » and « NOT ») and modifying the search to suit the different requirements of each database. Traditional systematic searches require prior knowledge of the topic of interest (the deeper the better). While deep knowledge and tailored search strings offer benefits, they also risk missing relevant articles on topics one did not think to include, and may limit the possibility of discoveries. Our software also allowed more exhaustive searching since it relied on full-text articles instead of only abstracts.

### Limitations

This version of the software is limited to databases that provide free APIs for open access to full-text articles. Databases often used for systematic reviews (e.g., Web of Science or Scopus), databases commonly used by social scientists (e.g., SocIndex, CINAHL, ATLA Religion, ProQuest services), and preprint servers for accessing non-peer reviewed literature (e.g. medRxiV, bioRxiV) did not provide suitable APIs. For example, Web of Science has a basic API but does not provide access to full-text articles. Some databases like Scopus or ProQuest provide a free basic API from which we could retrieve basic metadata but accessing more detailed information and full-texts data require a subscription. Medical preprint servers like medRxiv provide basic RSS feeds to obtain some metadata. Thus, our software is best for searching peer-reviewed, open-access medical literature, especially since most publications behind a paywall forbid text mining.

The Latent Dirichlet Allocation algorithm, like machine learning methods, has inherent limitations. The user must choose the number of topics; 10 is the default, but the ideal number of topics differs from corpus to corpus [24]. Our stepwise topic modelling approach mitigated this limitation. The LDA algorithm also performs badly on small corpora [24], so as our corpus was reduced, risk of incoherent topics increased. This is why we halted our iterations when the corpus shrank too much and analysed the parent corpus instead.

Our software only works on PDFs, but not yet on image-based PDFs. It also does not parse HTML-only publications. We are working on integrating optical character recognition (OCR) to translate images to texts. This limitation was reduced by our finding that image-based PDFs were usually less relevant, older articles.

Our broad search strategy (“HIV AND Malawi” anywhere in the text) retrieved many irrelevant articles that mentioned Malawi only in the references, so we intend to add the option to exclude references from the search.

Because the topics identified by the software sometimes overlapped, each article might fall under several topics, and each article was attributed only to the highest scoring topic, ambiguous or multiple attributions were ignored. Consequently, a relevant publication might have been classified in a topic of no interest, and thus was not incorporated in the final result.

Our software is intended to complement rather than replace systematic reviews. We have not yet compared our approach to a systematic review, and we expect we missed relevant articles unavailable through open-access. We also expect that our software missed some topics, while also finding new topics. As the number of open access articles and preprints increases [50], and as journals and preprint sites add APIs, we expect our software to become more useful. We plan to add more databases as we gain access.

We may have missed factors that influence HIV transmission in Malawi and it could be that broadening the search to “Malawi” would overcome this limitation and reveal possible interactions between social, political, economic and other factors that influence the course of the HIV epidemic, but which have never been studied in the context of HIV.

### HIV-specific discussion

Figure 3 shows the identified factors in a three-level diagram, after Kaufman et al. [51]. The 27 identified articles include 24 behavioural, social, and cultural factors of HIV infection. Compared to previous literature reviews on HIV-related factors observed worldwide [51-54], a few behavioural-related factors were not retrieved in our study. These include for example denial of HIV status, motivation and intention to change sexual behaviour, reactions to stress, physical and mental health status, outcome expectancies (i.e. anticipated consequences as a result of engaging in a specific behaviour) and empowerment. For social factors, relationship satisfaction and level of relationship commitment were also missing. Finally, we found no article discussing racism.

**Figure 3.**
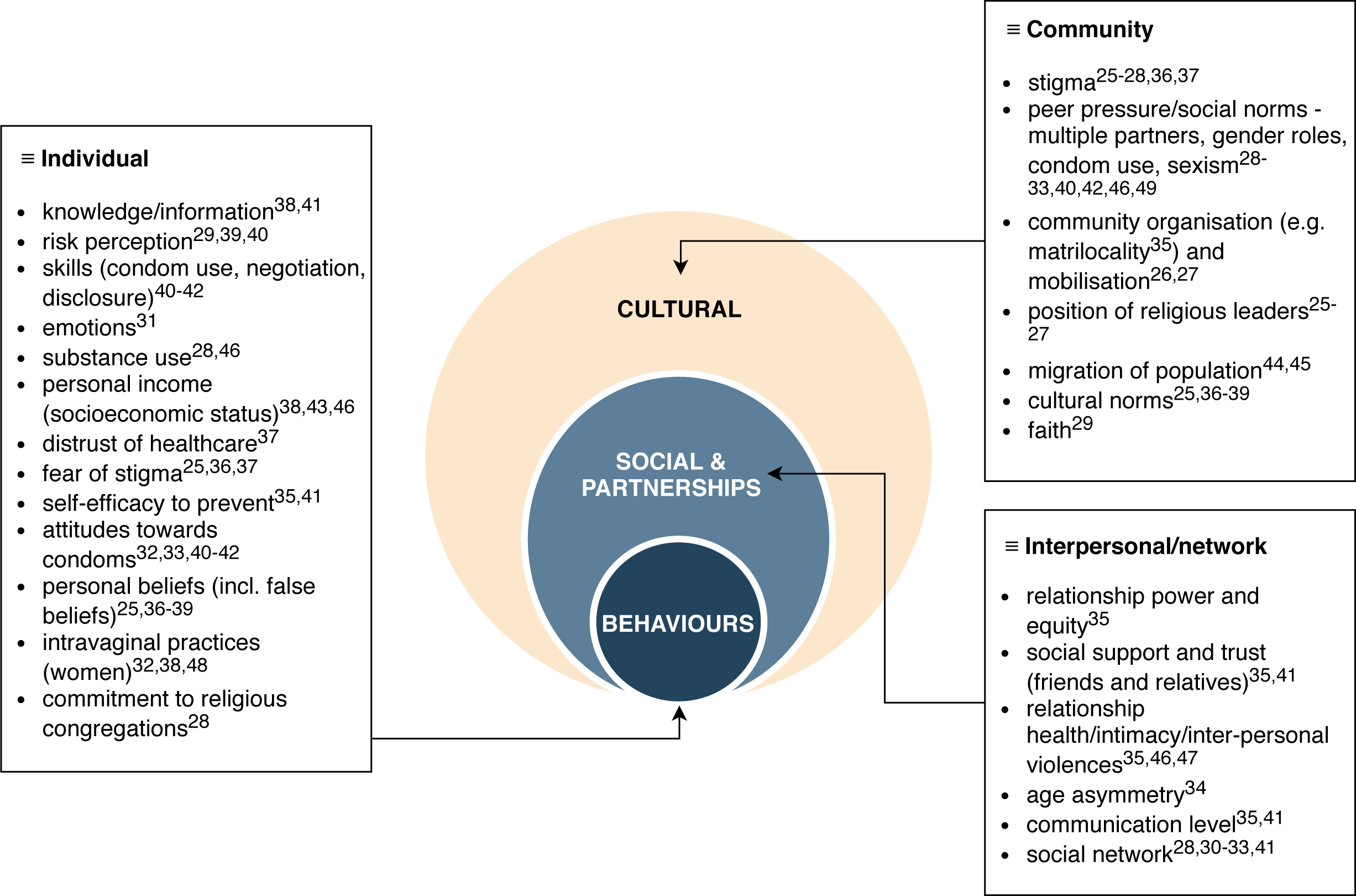
Summary of factors found within publications identified by the review.

The *2015-2020 National HIV Prevention Strategy* plan from the National AIDS Commission of Malawi targets specific HIV-related interventions at multiple levels for different population groups [55]. We compared the many behavioural, social, and cultural factors of HIV infection mentioned by the Prevention Strategy plan with the factors identified by our software. For the key populations of men having sex with men (MSM) and sex workers, reducing the number of partners, consistent use of condoms, targeted campaigns on HIV-testing and risk reduction, alcohol and substance abuse, positive health, and gender-based violence (GBV) prevention programmes are all behavioural interventions targetting risk factors that are present in our systematic review. Regarding the priority populations, additional interventions such as comprehensive sexuality and message on intergenerational sex for young women at risk, stop early marriage campaigns, female support for voluntary medical male circumcision (VMMC), HIV testing and counselling (HTC), and communication for couples are also addressing part of our behavioural list of HIV risk factors.

Behavioural factors, at an individual or a community level, that are not targeted by the Prevention Strategy plan for 2015-2020 were intravaginal practices. For the general population, distrust of healthcare, commitment to religious congregations, and the position of religious leaders are not addressed either. These are factors that should be considered when elaborating the new Malawi Prevention Strategy Plan for 2021-2026.

## Conclusions

From a set of articles limited by the existence of journal paywalls, our Python software quickly narrowed a set of over 16,000 articles to a small set of relevant articles. We identified socio-behavioural factors, including factors related to society and cultures like folk beliefs, theology and moral standards, that may influence the course of the HIV epidemic yet are rarely considered in the quantitative literature. Extending our approach to other countries could give researchers a more complete picture of the different drivers of the HIV epidemic in different settings and clarify the reasons for the spatial variability of HIV across sub-Saharan Africa.

## Data Availability

The data can be obtained directly in the list of publication databases mentioned in the manuscript.

